# Patterns and Trends of Antimicrobial Resistance of WHO Bacterial Priority Pathogens in Kenya: data from multi-site surveillance for the period 2021-2025

**DOI:** 10.64898/2026.08.14.26360438

**Authors:** Ali Kassim, Loice A. Ombajo, Josiah Njeru, Susan Githii, Cyrus Matheka, Joram Andrew, Edwin Otieno, Naomi Kariuki, Francis Kiigu, Victor Mburu, Julius Kiguru, Michael Kamau, Dorothy Kilonzo, Lilian Kutol, Denis Ndeto, Winnie Githinji, Grace Ndeda, Lydia Kabura, Wangu Githae, Peter Kiyondi, Rose Ndelema, Amos Walumbe, Morfat Okumu, Caroline Nzomo, Cosmus N. Ndeje, Christine Kinya, Christine N. Akoru, George Muchiri, Emmanuel Tanui, Christine M. Ngacha, Winnie Abuor, Duncan Nyukuri, Marybeth Maritim, Irungu Kamau

## Abstract

**Background:** Rising antimicrobial resistance (AMR) in the African region contributes to high morbidity and mortality. Continuous national AMR surveillance is critical in understanding the spread of AMR and informing policies on containment. We present results of national AMR surveillance in Kenya

**Methods:** Passive surveillance was prospectively conducted in 20 sites in Kenya between 2021 and 2025. Sites included national and sub-national level tertiary public and private hospital laboratories. Non-duplicate isolates of WHO priority Gram-negative and Gram-positive pathogens were included in this analysis. Bacterial isolates were identified using either conventional identification methods, Analytical Profile Index or automated systems while antimicrobial susceptibility testing was performed using the Kirby-Bauer disk diffusion method or automated systems and interpreted using the Clinical and Laboratory Standards Institute guidelines. The primary outcomes were the proportions of various priority bacteria isolated and the proportions resistant to commonly used antibiotics.

**Results:** Between 2021 and 2025, there were 15,124 priority pathogens isolated with 7,592 (50.2%) from urine, 5,430 (35.9%) from blood (35.9%), and 1,784 (11.8%) from respiratory specimens. *Escherichia coli* and *Klebsiella pneumoniae* accounted for 76.3% of the priority pathogens. Resistance to 3rd generation cephalosporins was 63.2% for *Escherichia coli* and 79.1% for *Klebsiella pneumoniae* for the period 2021 to 2025 while carbapenem-resistance was 30.4% for *Klebsiella pneumoniae* and 7.2% for *Escherichia coli*. Resistance to carbapenems by *Klebsiella pneumoniae* increased from 17.9% in 2021 to 35.9% in 2025 while Methicillin resistance in *Staphylococcus aureus* increased from 36.5% in 2021 to 56.4% in 2025.

**Conclusion:** Resistance to critical antibiotics is a significant problem in Kenya, with alarming rates of Methicillin Resistant *Staphylococcus aureus* and carbapenem resistant *Klebsiella pneumoniae*. Ugent and sustained infection prevention and control measures and appropriate antimicrobial stewardship activities should be instituted across all health facilities in the country. There is need for improved access to antibiotics with activity against these resistant pathogens.

## Introduction

Antimicrobial resistance (AMR) refers to a process through which micro-organisms such as bacteria, fungi and viruses, undergo evolutionary processes leading to resistance to antimicrobials commonly used in treatment of infections caused by such pathogens [1]. AMR is considered a global health concern with an associated 5 million deaths and 1.3 million deaths attributable to bacterial AMR in 2019. At a regional level, Sub-Saharan Africa (SSA) may have higher mortality attributable to AMR compared with other regions of the world [2], with mortality projected to rise if no concrete interventions are instituted [3].

A key factor in the emergence of AMR is the indiscriminate use of antibiotics in human, veterinary and agricultural settings. On average, the global antibiotic consumption rate in human health has risen from 9.8 Defined Daily Dose (DDD) per 1000 days in 2000 to 14.3 in 2018 [4]. Similarly, consumption of antibiotics in agriculture is postulated to increase by 30% by the year 2040 [5]. Increased use and misuse of antibiotics places micro-organisms under selective pressure to acquire adaptive mutations that confer resistance.

Holistic management of AMR should focus on three core areas: surveillance to detect shifting trends in AMR, infection prevention and control (IPC) measures, and improved access to diagnostic and treatment modalities for infections caused by resistant pathogens. Surveillance and control of AMR is best achieved by a targeted approach that prioritises pathogens that are of high public health concern. To this end, in 2017, the WHO developed and subsequently revised the bacterial priority pathogens list to highlight bacterial pathogens of public importance and to guide prioritisation of investment, research and AMR containment measures [6].

In 2014, WHO identified the African region as one of two regions without well-established surveillance system for antimicrobial resistance. National AMR surveillance coverage has gradually improved over the past 10 years. This is evidenced by the more than three-fold increase in the number of countries submitting AMR data to the WHO Global Antimicrobial Resistance and Use Surveillance System (GLASS) [7]. Kenya, for instance, has been submitting AMR data since 2021 and has progressively increased surveillance coverage. Despite this progress, completeness of AMR data was only documented in 54% of the 104 countries that submitted AMR data in 2023 [7]. We describe the national AMR trends from surveillance data in Kenya for the period between 2021 to 2025.

## Methodology

### Study Design and Setting

We prospectively collected data on bacterial isolates and antibiotic susceptibility tests of WHO priority pathogens isolated from WHO priority specimens. Clinical isolates from 20 sub-national and national referral public and private facilities (figure 1) from the year 2021 to 2025 were included. These facilities are located in areas where the majority of the Kenyan population live, with an estimated 50% of the Kenyan population living in the counties covered by the surveillance facilities [8]. During the study period, participating facilities were supported through procurement of microbiology equipment, laboratory reagents and supplies, strengthening of antimicrobial stewardship committees to improve the clinical-laboratory interphase, and training and mentorship of laboratory personnel on processing and reporting of bacterial isolates.

**Fig 1:**
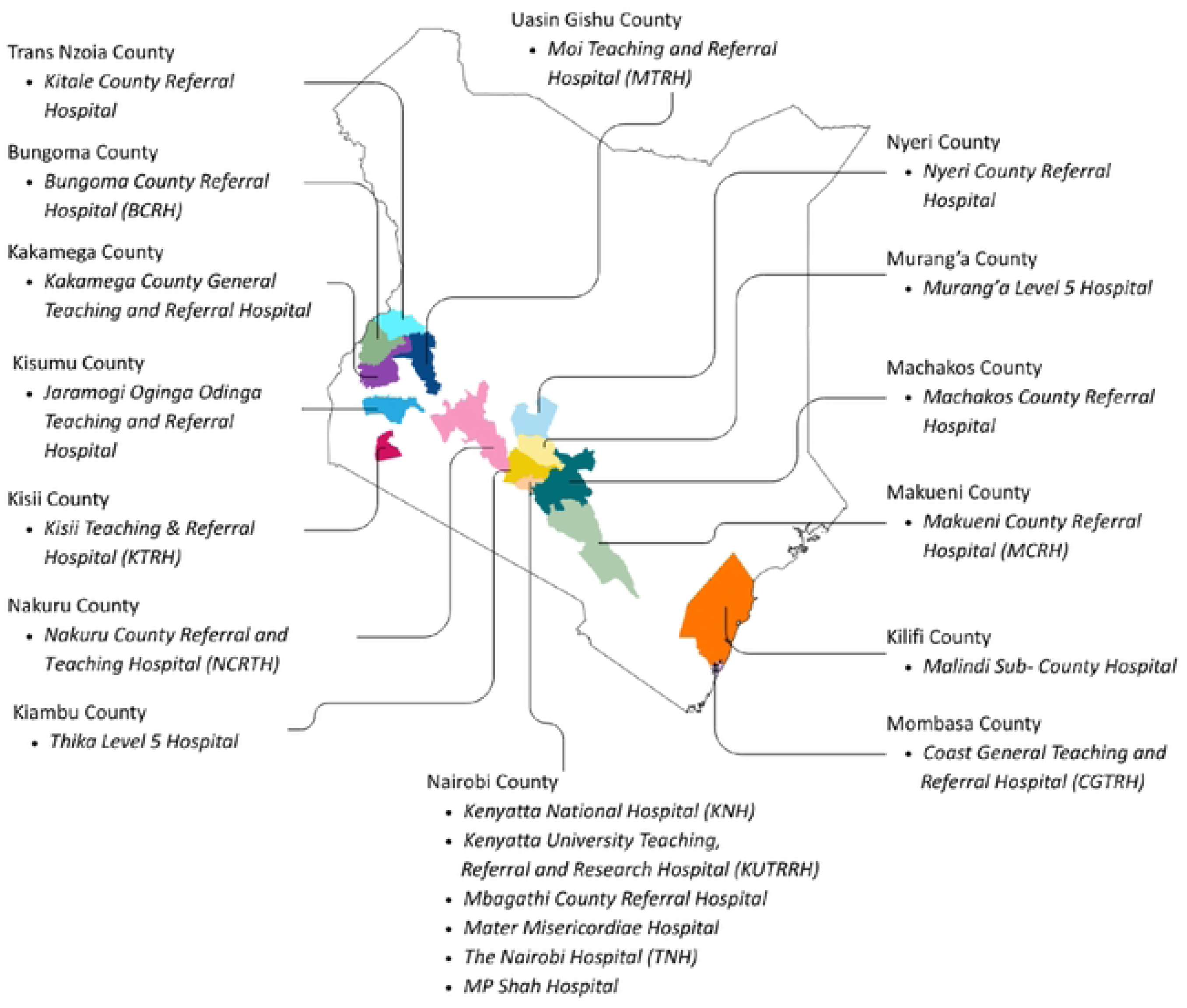
Map of Kenya showing the distribution of the AMR surveillance sites.

Microbiological specimens in these facilities were processed following facility based standard operating procedures aligned with Clinical Laboratory Standards Institute (CLSI) and WHO guidelines. Bacterial isolates were identified using either conventional identification methods, API systems or automated systems such as Vitek® 2 system (bioMérieux, Marcy-l’Étoile, France), BD Phoenix^TM^ M50 (Becton Dickinson, Sparks, MD, USA) and MALDI-TOF MS systems (Bruker Daltonics, Bremen, Germany) depending on the capacity of the facility. Antimicrobial susceptibility testing was performed using the Kirby-Bauer disk diffusion method and/or automated antibiotic susceptibility testing (AST) systems. Choice of antibiotics and interpretation of susceptibility results was guided by the latest CLSI M100 guideline. Majority of the laboratories are accredited to ISO 15189:2012. Quality Assurance was done using reference American Type Culture Collection (ATCC) strains as well as participation in external quality assessment (EQA) programs coordinated by the National Public Health Laboratory (NPHL) and the External Quality Assessment for AMR Testing for Africa (EQuAFRICA). Each facility submitted the microbiological data to the Ministry of Health-NPHL’s Central Data Warehouse as part of the National surveillance system.

Priority samples and isolates were aligned to the WHO GLASS guidance on AMR surveillance and the Kenyan AMR surveillance strategy. Antimicrobial drugs were selected depending on CLSI tiered selection of antibiotics for different pathogens and their clinical use in the different priority samples. The AMR surveillance, laboratory capacity building including training and supply of consumables, EQA programme, data cleaning and analysis were supported by the Fleming Fund Country Grant to Kenya. Data analysis and visualisation was supported by the Center for Epidemiological Modelling and Analysis (CEMA) at the University of Nairobi (UON).

This work was conducted as part of national surveillance, did not involve any direct patient identifiers and did not require ethical approval.

### Data Management and Analysis

Data from all participating surveillance sites were continuously submitted to the national central data warehouse (CDW) hosted at the National Public Health Laboratory (NPHL). Submissions were received in multiple formats, including WHONET exports, Laboratory Information Management System (LIMS) outputs, and Vitek® 2 instrument files generated by facility laboratories. Data received was reviewed monthly for completeness. Biannually, data was cleaned by a working group comprising of the surveillance site microbiologists, microbiology technologists, infectious disease physicians, clinical pharmacists and data scientists. The data was assessed for completeness, relevance, and adherence to the correct priority pathogens - antibiotics combinations. To ensure the consistency, reliability, and quality of the compiled dataset, annual data review and analysis workshops were conducted. These workshops also served as capacity-building platforms for equipping AMR focal persons with the skills and contextual understanding needed to actively contribute to the national data harmonisation process.

The dataset was cleaned in accordance with Clinical and Laboratory Standards Institute (CLSI) guidelines which addressed the following two primary areas;

#### De-duplication of isolates

Patients are frequently sampled on multiple occasions - either for diagnostic purposes or to monitor therapeutic response - and because those harbouring resistant organisms were disproportionately more likely to yield repeated cultures, uncleaned data carried an inherent risk of measurement bias. To address this, duplicate isolates were identified and removed in accordance with GLASS recommendations. For each 12-month surveillance period, only the first isolate per patient, per surveyed specimen type, and per surveyed pathogen was retained for analysis. In instances of insufficient data, unique identifiers were constructed from available demographic data to support accurate de-duplication.

#### Standardisation of antimicrobial susceptibility testing results

All AST results were reviewed and re-coded against CLSI breakpoints to ensure comparability across sites and surveillance periods. Records containing incomplete or invalid AST data were flagged during the data review workshops and resolved through direct consultation with the originating facility laboratories.

### Descriptive Analysis

Following data cleaning and standardisation, descriptive analyses were conducted using R statistical software. This enabled efficient processing of the cleaned dataset and the systematic generation of summaries consistent with international AMR surveillance standards. Outputs included pathogen-specific resistance proportions, specimen-type distributions, and temporal resistance trends across participating surveillance sites.

## Results

The most common specimens processed by participating laboratories were urine, blood and respiratory specimens accounting for 97.9% of priority specimens. 50.2% of the priority pathogens reported during the surveillance period 2021 to 2025 were isolated from urine, 35.9% from blood and 11.8% from lower respiratory specimens.

### Distribution of priority pathogens over the surveillance period

There were 15,124 priority pathogens isolated between 2021 to 2025, with the number of pathogens rising from 1,539 to 5,570 over this period, representing a 261% increase. *Escherichia coli* and *Klebsiella pneumoniae* accounted for 76.3% of priority pathogens The table 1 below shows the distribution of priority pathogens over the surveillance period 2021 to 2025.

**Table 1:**
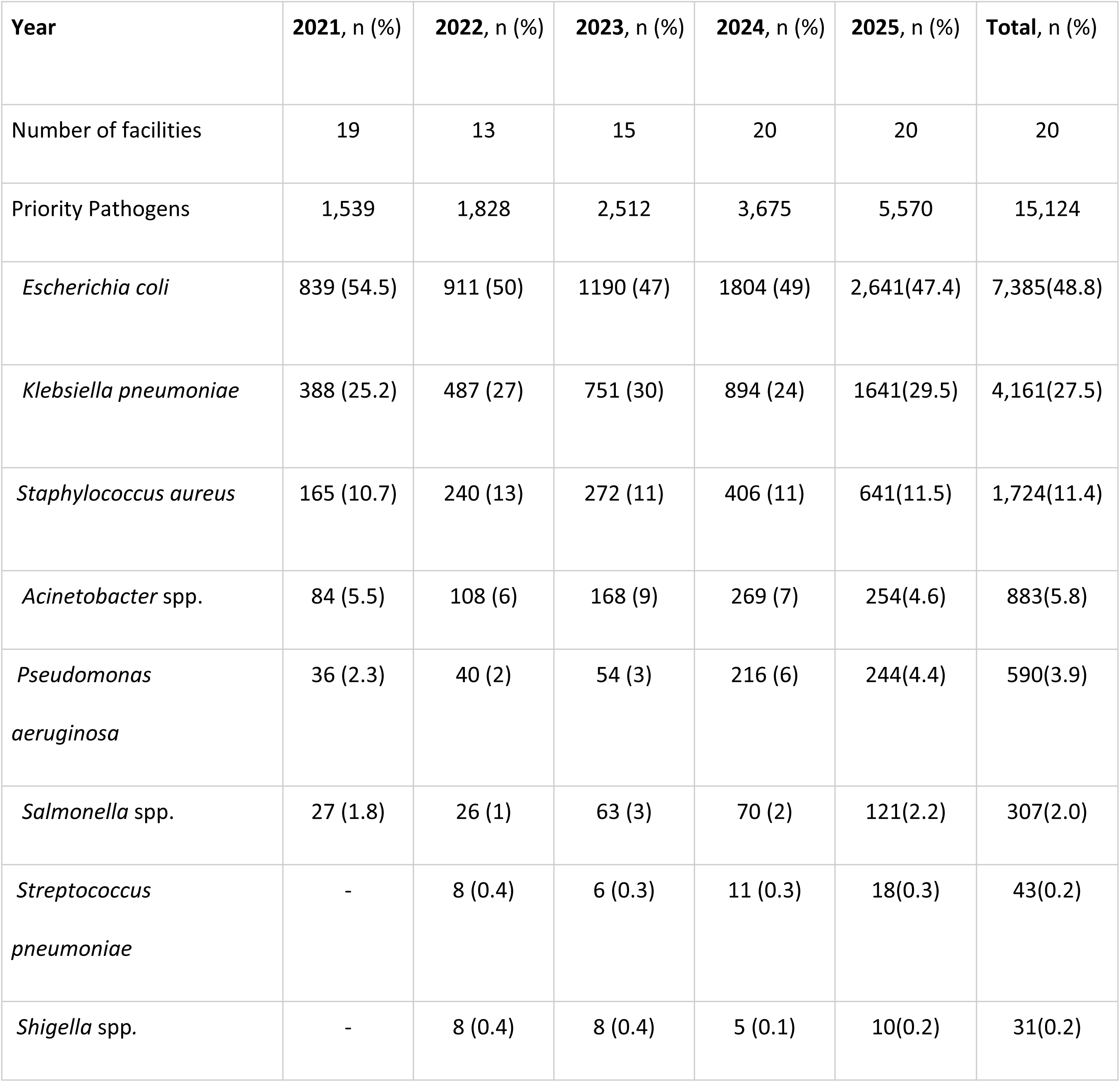
Distribution of priority pathogens reported in the surveillance period 2021 to 2025.

### Distribution of priority pathogens by specimen types

The most common priority pathogen isolated from urine was *Escherichia coli* accounting for 76.4% while *Klebsiella pneumoniae* accounted for 23.6%. The most common isolates from blood were *Klebsiella pneumoniae* at 32.5%, *Staphylococcus aureus* at 26.9% and *Escherichia coli* at 24.5%. *Acinetobacter* species, *Pseudomonas aeruginosa* and *Salmonella* species accounted for 9.5%, 3.6% and 2.6% of blood isolates respectively. *Klebsiella pneumoniae* accounted for the majority of priority pathogens isolated from lower respiratory specimens at 32.7%, followed by *Pseudomonas aeruginosa* at 21.0%, *Acinetobacter* species at 18.8%, *Escherichia coli* at 13.3% and *Staphylococcus aureus* at 12.9% (Table 2).

**Table 2:** Distribution of Priority Pathogens by priority specimen types over the surveillance period.

| **Priority Pathogen** | **Urine, %**<br>**(n=7,590)** | **Blood, %**<br>**(n=5430)** | **Lower Resp.**<br>**specimens, %**<br>**(n=1,783)** | **Stool, %**<br>**(n=190)** | **Cerebrospinal**<br>**fluid, %**<br>**(n=131)** |
| --- | --- | --- | --- | --- | --- |
| *Escherichia coli* | 76.4 | 24.5 | 13.3 | - | 14.5 |
| *Klebsiella pneumoniae* | 23.6 | 32.5 | 32.7 | - | 16.0 |
| *Staphylococcus aureus* | - | 26.9 | 12.9 | - | 27.5 |
| *Acinetobacter* spp. | - | 9.5 | 18.8 | - | 24.4 |
| *Pseudomonas aeruginosa* | - | 3.6 | 21.0 | - | 14.5 |
| *Salmonella* spp. (non-typhoidal) | - | 2.6 | - | 83.2 | 2.3 |
| *Streptococcus pneumoniae* | - | 0.3 | 1.3 | - | 0.8 |
| *Shigella* spp. | - | - | - | 16.3 | - |
| *Salmonella* Typhi | - | 0.04 | - | 0.5 | - |

### AMR patterns in Kenya for the period 2021 to 2025

The overall rate of resistance to third generation cephalosporins was 64% in *Escherichia coli* and 79% in *Klebsiella pneumoniae* for the period between 2021 to 2025. Overall Carbapenem resistance in *Klebsiella pneumoniae* during this period was 30% while that in *Escherichia coli* was 7%. In 2025, the overall national rate of carbapenem resistance in *Klebsiella pneumoniae* was 36% with notable regional variation. Figure 2 below shows the AMR patterns for the Gram-negative priority pathogens over the 2021 to 2025 period. Figure 3 shows the regional spread of carbapenem resistant *Klebsiella pneumoniae*. The overall MRSA rate for the period between 2021 to 2025 was 47%, with a rate of 56% in 2025 alone.

**Fig 2:**
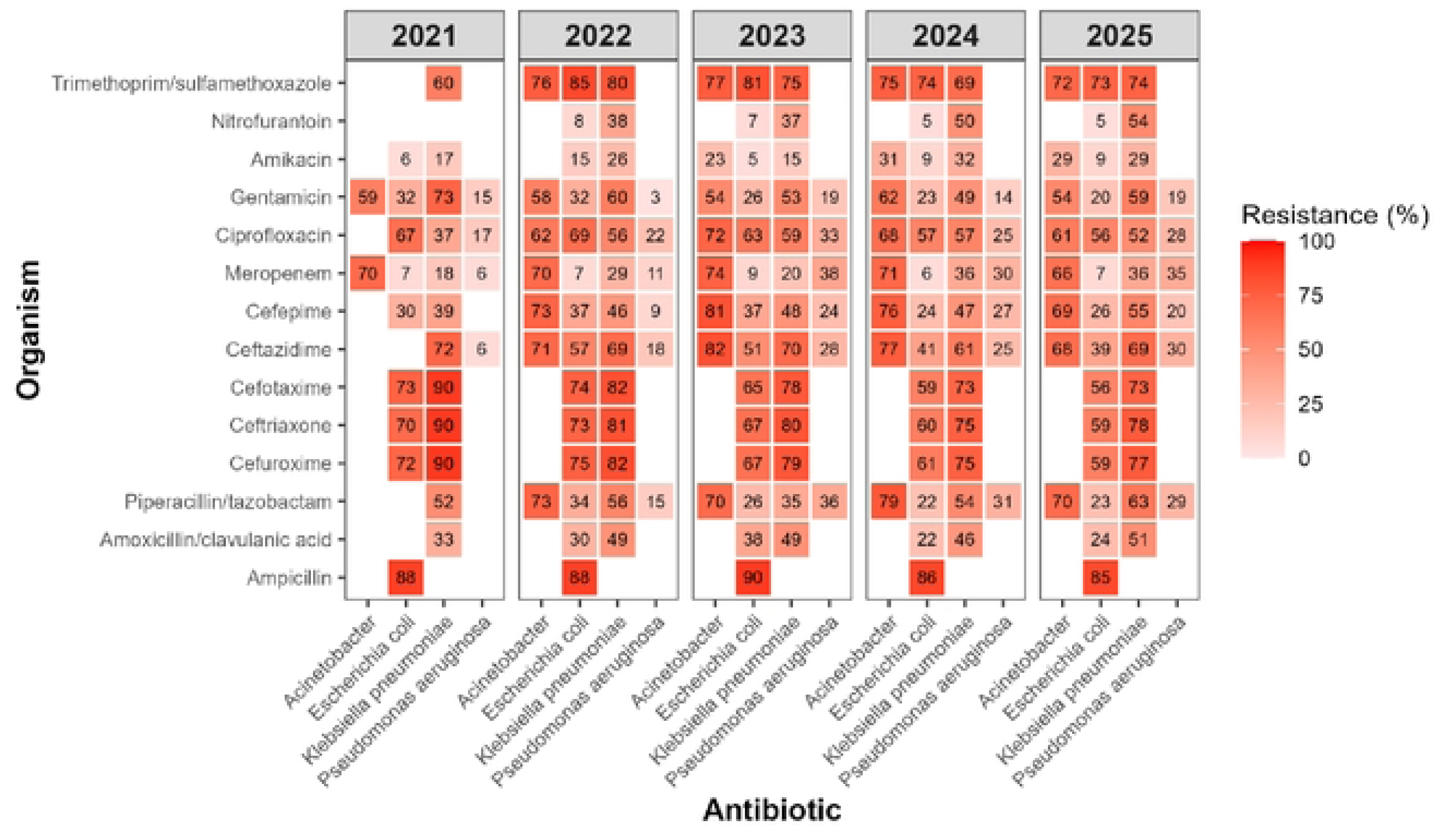
AMR Patterns for the Gram-Negative Priority Pathogens over the surveillance period 2021 to 2025.

**Fig 3:**
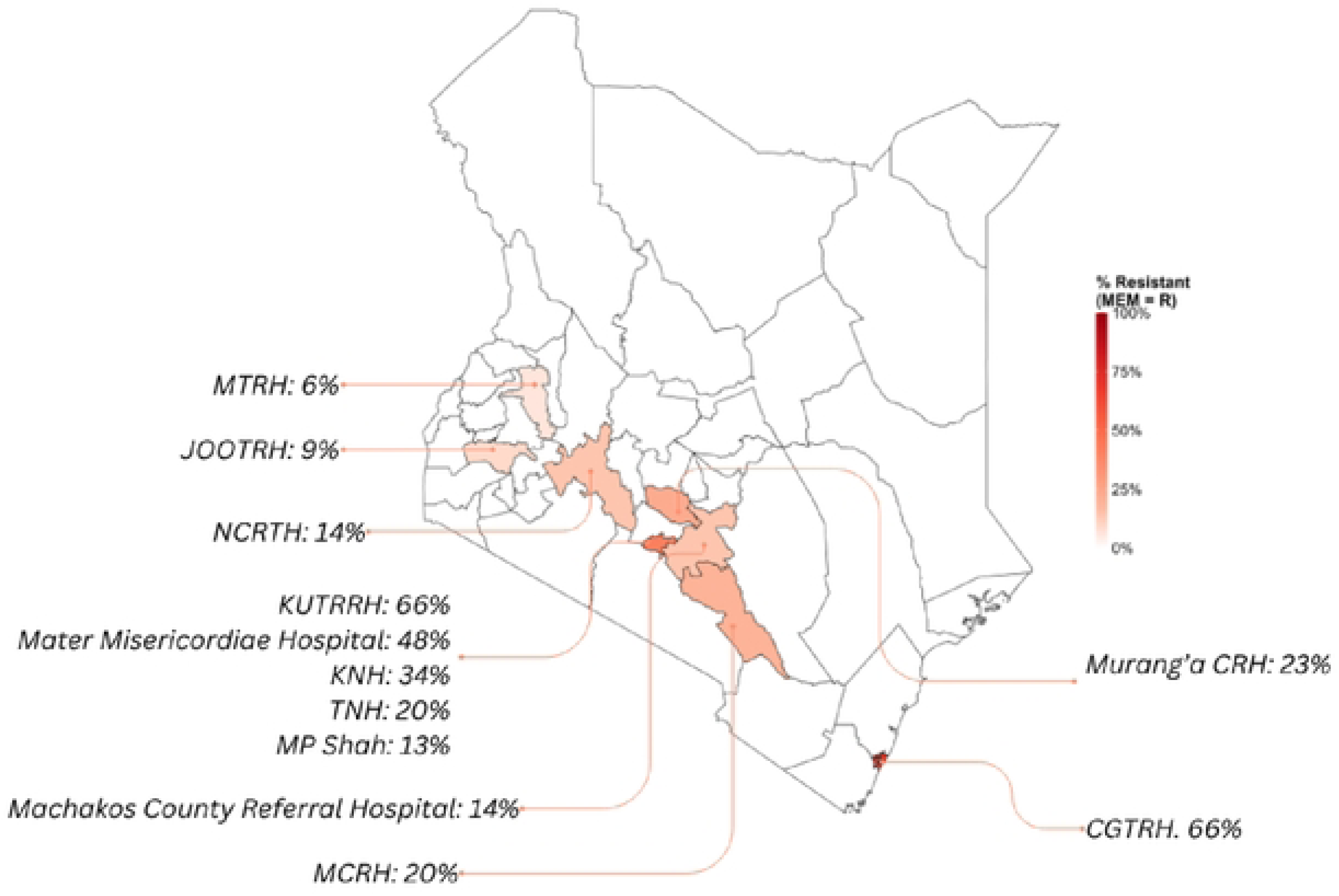
Map of Kenya showing the distribution of *Klebsiella pneumoniae* resistance to Meropenem 2025.

### Trends of Antimicrobial Resistance in Gram Negative bacteria from 2021 to 2025

*Escherichia coli* resistance to cephalosporins, ciprofloxacin, gentamicin and trimethoprim-sulfamethoxazole reduced slightly while resistance to meropenem, piperacillin-tazobactam, amoxicillin-clavulanate, ampicillin, nitrofurantoin and amikacin were steady or with slight fluctuations over the surveillance period (figure 4). *Klebsiella pneumoniae* showed increased resistance to Amikacin, Meropenem, Piperacillin-tazobactam and nitrofurantoin with slight reductions in resistance for the other antibiotics (figure 5). The rate of resistance of *Acinetobacter* species to most antibiotics was stable during the assessment period (figure 6), while resistance of *Pseudomonas aeruginosa* increased between 2021-2023 and remained stable between 2023-2025 (figure 7)

**Fig 4:**
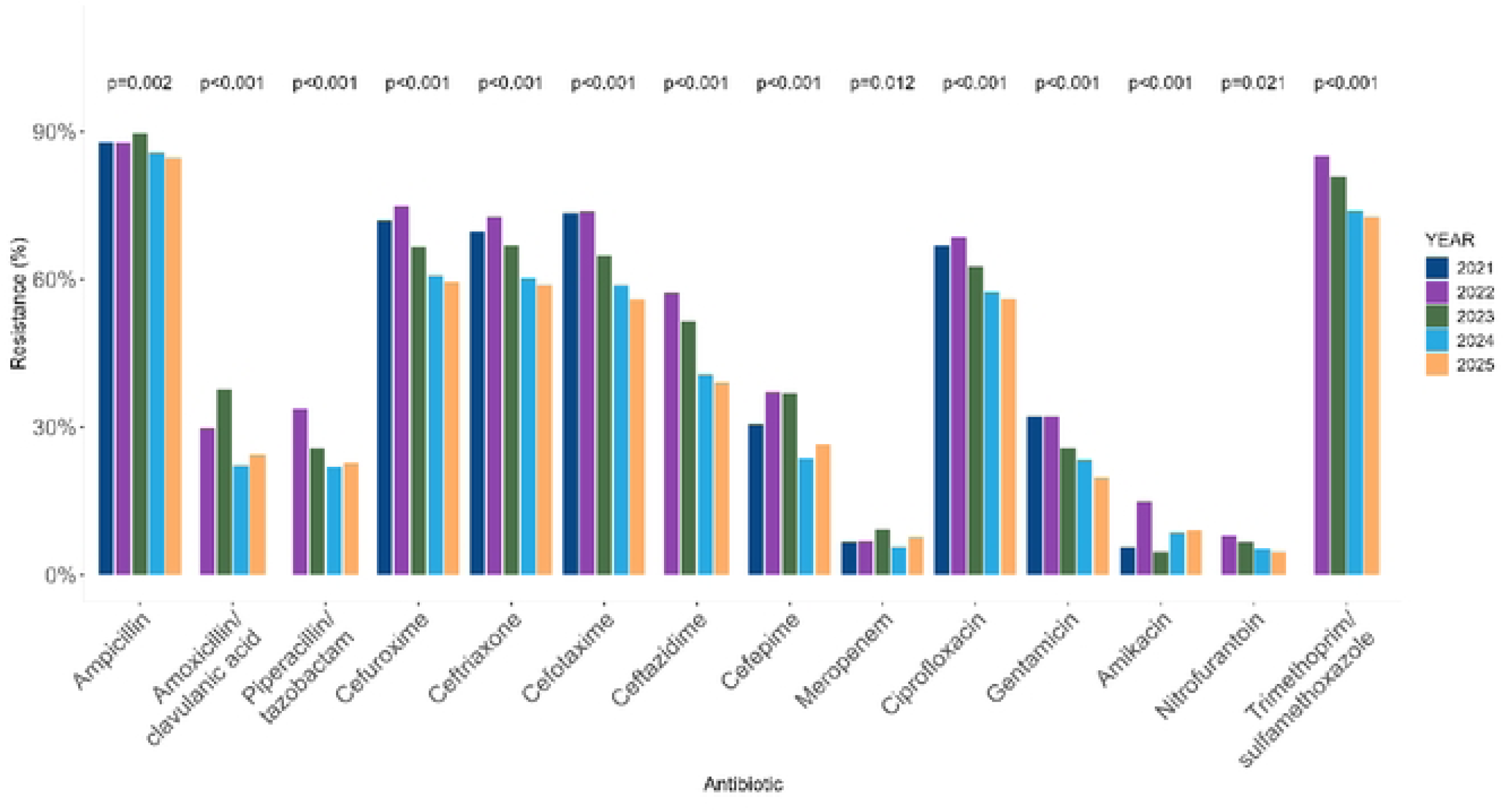
Trend analysis of AMR in *Escherichia coli* to the commonest antibiotics between 2021 and 2025.

**Fig 5:**
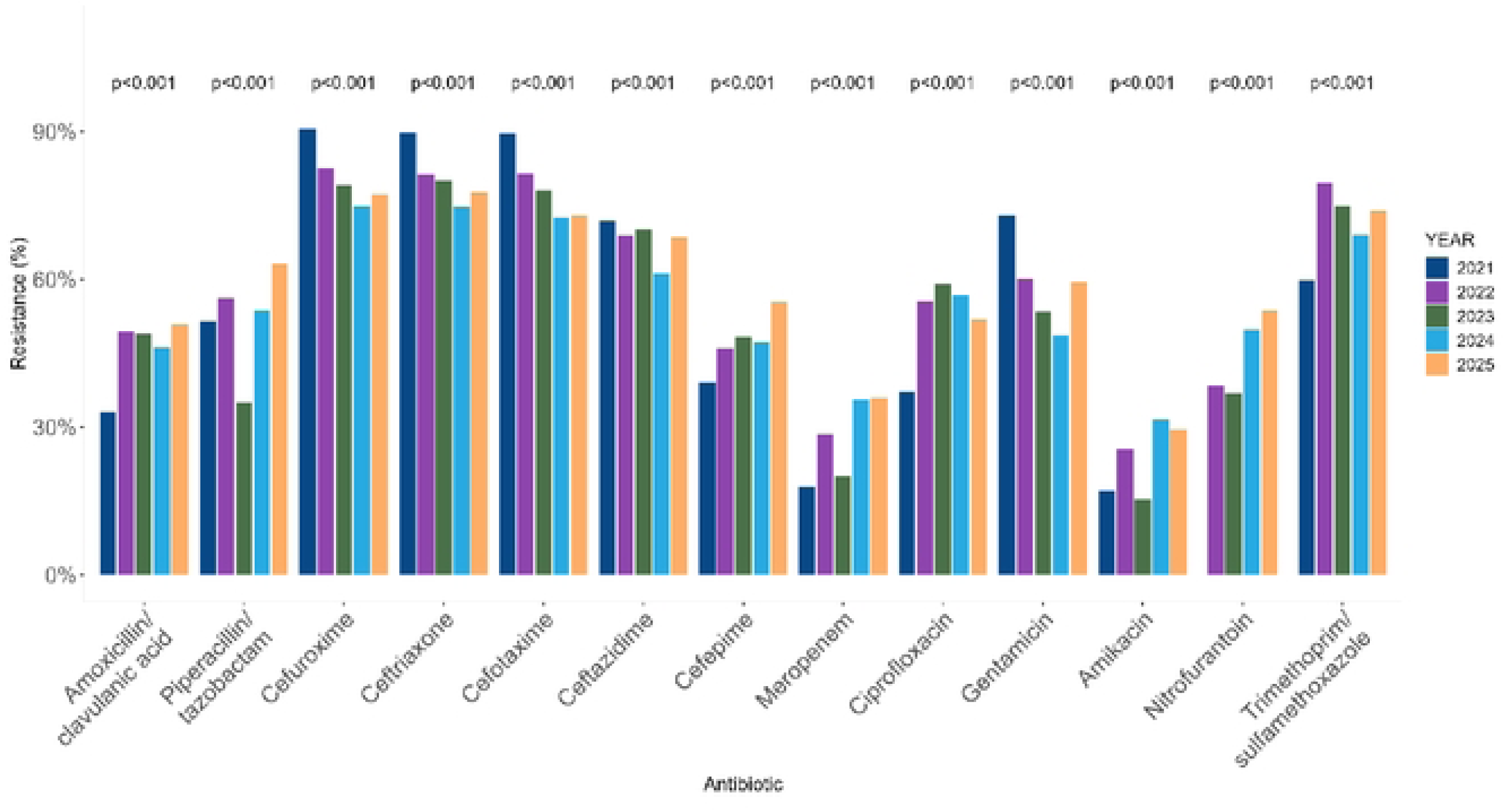
Trend analysis of AMR in Klebsiella pneumoniae to the commonest antibiotics between 2021 and 2025.

**Fig 6:**
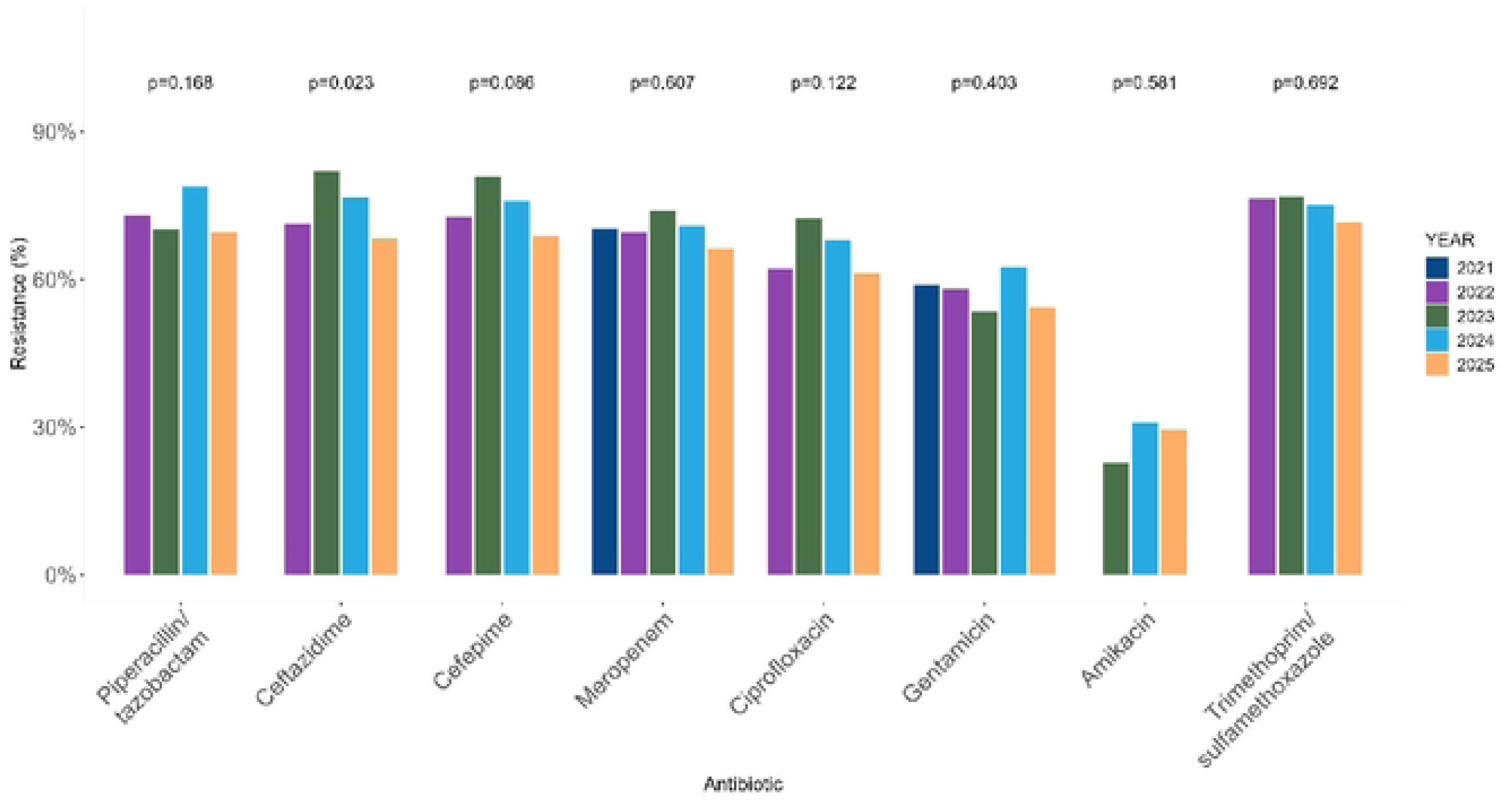
Trend analysis of AMR in Acinetobacter spp. to the commonest antibiotics between 2021 and 2025.

**Fig 7:**
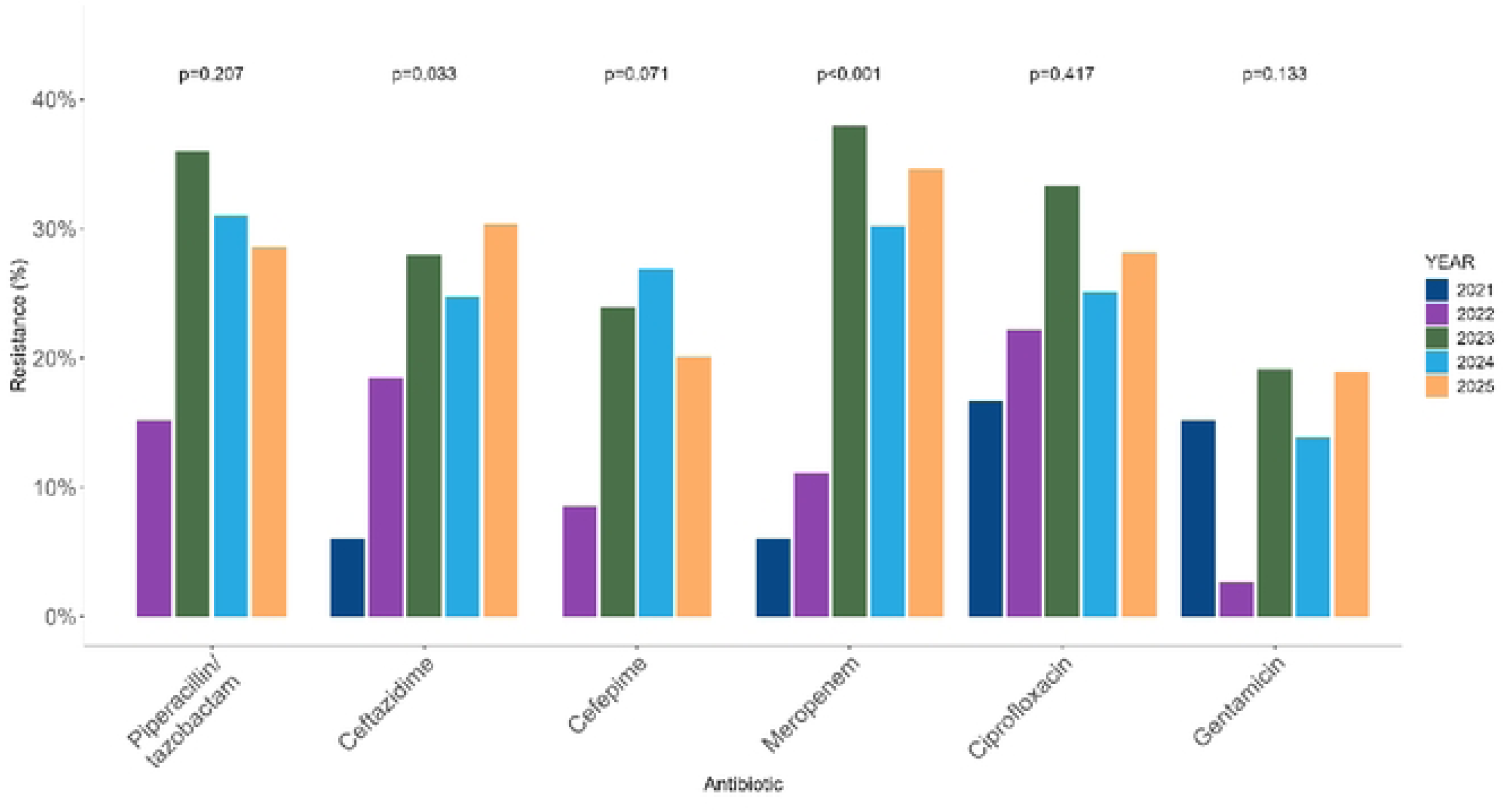
Trend analysis of AMR in Pseudomonas aeruginosa to the commonest antibiotics between 2021 and 2025.

**Fig 8:**
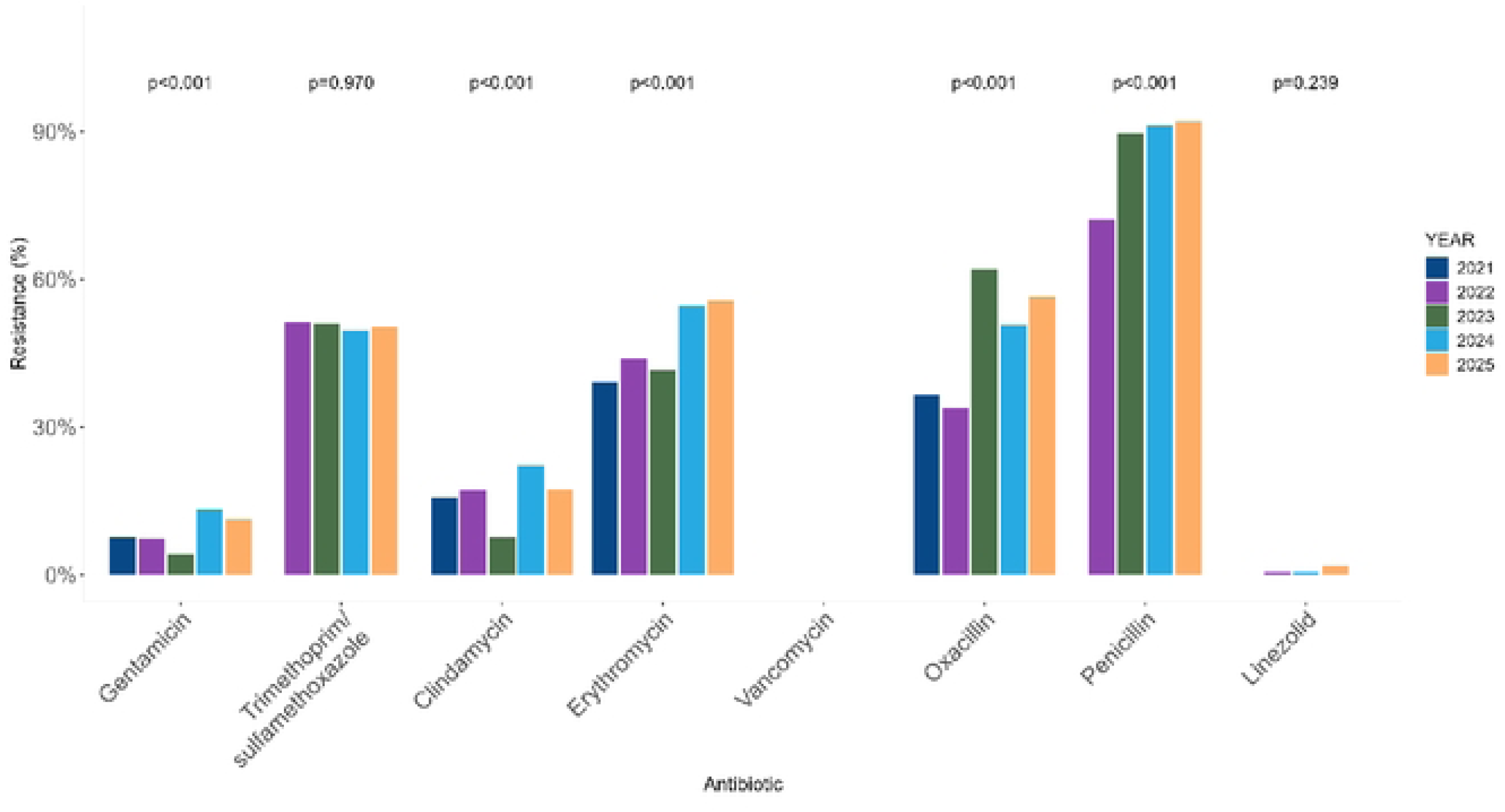
Trend analysis of AMR in Staphylococcus aureus to the commonest antibiotics between 2021 and 2025.

### Trends of AMR of Gram-positive organisms in Kenya from 2021 to 2025

The overall rate of MRSA was high in all facilities and increased from 37% in 2021 to 56% in 2025. Resistance to trimethoprim-sulfamethoxazole was 50% and did not change over the surveillance period. Clindamycin resistance increased from 16% in 2021 to 22% in 2024 with a reduction to 17% in 2025. Resistance to macrolides, represented by erythromycin, increased from 39% to 56% over the surveillance period. There was no confirmed resistance to Vancomycin (figure 7)

## Discussion

Support for strengthening of AMR surveillance systems in Kenya led to a 261% increase in priority pathogens reported from 1,539 in 2021 to 5,570 in 2025. Surveillance data in low and middle-income countries, particularly in Africa, has been poor, marked by significant fragmentation, lack of consistency and low quality [9]. This is due to the significantly low laboratory infrastructure and capacity in many of these settings. [7,10] The increase in surveillance data demonstrated in this study is an indication of the progress that can be achieved with improved funding, better governance and enhanced human resource capacity in low resource settings. Resources such as those provided by the Fleming Fund can lead to significant improvements in microbiology laboratory capacity but it is incumbent upon governments to sustain these gains.

*Escherichia coli* and *Klebsiella pneumoniae* accounted for 48.8% and 27.5% of the overall priority pathogens and 76.4% and 23.6% of urinary priority pathogens respectively. This is consistent with other surveillance data from Africa showing *Escherichia coli* and *Klebsiella pneumoniae* as the commonest pathogens [11,12]. *Klebsiella pneumoniae, Staphylococcus aureus* and *Escherichia coli* together accounted for over 80% of pathogens isolated from blood cultures with rates of 32.5%, 26.9% and 24.5% respectively. More recent studies across Africa and globally show similar patterns in contrast with results from older studies in which *Salmonella enterica* and *Streptococcus pneumoniae* predominated [13,14].

*Klebsiella pneumoniae*, *Acinetobacter spp*. and *Pseudomonas aeruginosa* were the commonest isolates from respiratory specimens, particularly tracheal aspirates from patients in critical care units. These patterns may reflect colonisation of endotracheal tubes as well as increasing prevalence of ventilator-associated pneumonia in critical care units as described in other studies [15–17].

Cerebrospinal fluid (CSF) and stool specimens with priority pathogens accounted for about 1% each. The low rates of priority pathogens from these specimens could be due to low utilisation of the microbiology laboratory in the setting of meningitis and diarrhoea, specimen collection after initiation of antibiotics, difficulty in specimen collection and a lower prevalence of these infections. The commonest pathogens isolated from CSF include *Staphylococcus aureus* at 27.5%, *Acinetobacter* species at 24.4%, *Klebsiella pneumoniae* at 16.0%, *Escherichia coli* at 14.5 and *Pseudomonas aeruginosa* at 14.5%. The isolation of these atypical pathogens may represent a selection bias with specimen more likely to be collected from patients with longer hospital stay and in those with hospital rather than community acquired infections. Non-typhoidal *Salmonella* species and *Streptococcus pneumoniae* were rare at 2.3% and 0.8% respectively. The low number of *Streptococcus pneumoniae* isolated from blood, respiratory and CSF samples may be due to widespread uptake of pneumococcal vaccines in the paediatric population in Kenya. This has been documented in other similar contexts [18].

Of the enteric pathogens, non-typhoidal *Salmonella* constituted 83.2% of isolates, *Shigella* spp. 16.3%, and *Salmonella* Typhi isolation was low at 0.5%. In contrast to studies showing equal or higher rates of *Shigella*, *Salmonella* species were the more predominant enteric pathogen in this study [19]. In contrast to previous data suggesting *Salmonella* Typhi as a common and major cause of bacterial febrile disease [20], the rate of isolation of *Salmonella* Typhi from both stool and blood specimens was very low at all the surveillance sites. All surveillance data was collected prior to roll-out of the *Salmonella* Typhi vaccine in the country.

We found that *Escherichia coli* and *Klebsiella pneumoniae* had high rates of resistance to ceftriaxone ranging from 58.8% to 72.8% and 74.6% to 89.9% respectively over the surveillance period. This is in keeping with findings from Kenya and the wider African region. Bwanali *et al* reported resistance of Gram-negative organisms to ceftriaxone of 63.0% to 72.4% in Malawi [9]. Maina *et al* reported resistance of *Escherichia coli* to third generation cephalosporins at 79% in a private Kenyan hospital [21]. High antibiotic use, particularly of third generation cephalosporins such as ceftriaxone has been reported in Kenyan hospitals [22]. The odds of colonisation with extended-spectrum cephalosporin resistant and carbapenem-resistant enterobacteriacae in Kenya has also been shown to be higher with ceftriaxone use [23].

We demonstrate increasing resistance by *Klebsiella pneumoniae* to amikacin, meropenem, piperacillin-tazobactam and nitrofurantoin. Meropenem resistance in *Klebsiella pneumoniae* was significantly high ranging from 17.9% to 35.9% with resistance of 5.7% to 9.3% in *Escherichia coli*. Yakobi *et al*, in a systematic review and meta-analysis of surveillance studies from a South African population, reported high meropenem resistance of 31.7% in *Klebsiella pneumoniae* [24]. This rising rate of carbapenem resistance in *Klebsiella pneumoniae* in Africa and globally has been shown in several studies, including studies in neonatal and critical care patient populations [25,26]. Resistance to carbapenems is particularly concerning for Kenya and other resource limited settings as carbapenems are often the last resort antibiotics available with limited or non-existent access to newer reserve antibiotics.

*Acinetobacter* spp. isolates were noted to exhibit resistance to most antibiotics including third and fourth generation cephalosporins, meropenem, ciprofloxacin, piperacillin-tazobactam and trimethoprim-sulfamethoxazole. The rates of resistance were relatively unchanged over the 4-year period. Amikacin demonstrated the highest activity against *Acinetobacter* spp. isolates. These findings are comparable to global trend and susceptibility profiles from the African region [7]. The prevalence of carbapenem resistant *Acinetobacter baumannii* (CRAB) ranged from 66-74%, highlighting the challenge in treating *Acinetobacter baumannii* infections. This is similar to rates reported in other studies from Kenya, with CRAB rates of 83% [27]. However, some studies from the African region reported lower CRAB rates of 33-44% [28]. *Pseudomonas aeruginosa* isolates exhibited higher resistance rates against Piperacillin-Tazobactam and meropenem (30-31%). The prevalence of carbapenem resistant *Pseudomonas aeruginosa* was comparable to global rates of 33% resistance to imipenem) and 23% resistance to meropenem) [29].

MRSA rates were high and increased over the surveillance period from 37% in 2021 to 56% in 2025. In a Malawian study, the MRSA rate was reported to increase from 66.7% in 2020 to 84.2% in 2023 with a drop to 75.0% in 2024 [11]. There were no confirmed Vancomycin resistant *Staphylococcus aureus* (VRSA) isolates in this study. However, studies across Africa before 2020 showed an average VRSA rate of 2.5% with more recent studies showing rising rates [30,31].

The rise in resistant pathogens, particularly resistance to carbapenems by Klebsiella *pneumoniae* underscores the urgent need for improvement of infection control, particularly in hospital settings, to contain spread [32]. It also highlights the critical need for the development of vaccines against Klebsiella *pneumoniae*, the need for improved antimicrobial stewardship and the very pressing need for improved access to antibiotics with activity agains carbapenem resistant enterobacteriacae. Efforts by governments in resource limited settings to improve multi-sectoral and community engagement for a one health approach to infection prevention and reduction of AMR [32] as well as allocation of resources for surveillance is crucial.

The main limitation of this study is the fact that only national referral and higher-level county facilities were involved in this surveillance. This is due to the fact that lower-level facilities often do not have sufficient microbiology capacity to produce quality data. Data from higher level facilities may be skewed towards demonstrating higher levels of resistance as patients in these facilities are often sicker, may have been exposed to multiple hospitalisations and courses of antibiotics all of which promote a profile of resistance.

## Conclusion

The study reveals high rates of carbapenem resistant *Klebsiella pneumonia and* Methicillin Resistant *Staphylococcus aureus* in Kenya. This underscores the need to urgently put in measures to control inappropriate antibiotic use, strengthen infection prevention and control and support ongoing surveillance.

## Data Availability

Replication data for this study is available with publication at the Harvard Dataverse by request to the Principal Investigator (L.A.O) based on an approved proposal, at https://doi.org/10.7910/DVN/HKA3EF. Data on clinical isolates and the data dictionary defining all fields in the set are included.

## Acknowledgements

We acknowledge logistic support from the Kenyan Ministry of Health through the National Antimicrobial Stewardship Interagency Committee, county governments, leadership of the hospitals that provided AMR data, International Livestock Research Institute and Mott McDonald

